# Incidence and Prevalence of Hidradenitis Suppurativa: A Systematic Review and Meta-analysis

**DOI:** 10.1101/19012559

**Authors:** N. Gill, R. Gniadecki

## Abstract

**Background:** Hidradenitis is a chronic relapsing follicular occlusive disease with a widely variable reported prevalence. The exact prevalence and incidence of HS is unknown.

**Objectives:** To perform a systematic review and meta-analysis of the published literature to estimate the global incidence and prevalence of HS.

**Methods:** Literature searches were performed on Medline, Embase, and Pubmed to identify studies reporting incidence and/or prevalence of HS. Pooled estimates of prevalence and incidence were calculated with a meta-analysis of proportions.

**Results:** In total, 12 studies were included (Australia, Brazil, Denmark, France, Germany, Ireland, Israel, UK, USA) comprising a total population of 53,805,690. Eleven studies reported prevalence. The pooled proportion of individuals in the general population with HS was 0.36% (95% CI 0.21 - 0.56). Self-reported HS gave a higher prevalence estimate than clinician-diagnosed HS. HS is more prevalent in women. Average annual incidence of HS was 28.5 cases/100,000 (95% CI 26.8 – 30.1).

**Conclusions:** We estimated the global prevalence of HS to be 0.36% with 3:2 female predominance and average annual incidence to be 28.5 cases/100,000.

## Introduction

Hidradenitis suppurativa (HS) is a follicular occlusive disease presenting with recurrent abscesses, scarring, and sinus tract formation in the apocrine gland bearing regions. The cause of the perifollicular inflammation in HS is largely unknown^1^ but may be related to abnormalities in the keratinocyte signaling^2,3^, aberrant immune response to normal skin flora^4^ or a biofilm-driven process causing chronic inflammation of lesions.^5^ A diagnosis of HS is made clinically based on the presence of typical lesions (recurrent inflamed nodules progressing to abscesses, sinus tracts, and/or fibrotic scars) in the often bilateral distribution in the axillae, groin, and inframammary areas.^6^ The Hurley clinical staging system^7^ is used to categorize disease severity into three stages: stage I (abscess formation without sinus tracts and scarring), stage II (recurrent single or multiple widely separated abscesses with sinus tracts and scarring), stage III (multiple interconnected sinus tracts and abscesses).

Reported prevalence varies widely from 0.05 – 4.10%^1^ with a female predominance. Symptom onset typically occurs between puberty and age 40. HS is notoriously difficult to treat with recurrent painful flares resulting in impaired quality of life (QoL)^8^ and significant economic strain on the healthcare system through repeated emergency visits and hospitalizations.^9^ QoL is low even when compared to other chronic inflammatory skin diseases such as psoriasis.^10^ Newer treatments, such as adalimumab, have been shown to control disease^11^ and achieve significant cost-savings through containment of exacerbations.^12^ Continued research and innovation in the area of HS treatment requires knowledge of incidence and prevalence. The aim of the present study to summarize and evaluate the published estimates for the global prevalence and incidence of HS.

## Materials and Methods

### Search strategy

We searched the Ovid MEDLINE, Ovid Embase, and Pubmed databases for papers reporting the prevalence and/or incidence of HS. Studies were considered for inclusion if they met the predefined inclusion criteria: reported data required to calculate the prevalence and/or incidence of HS in the general population, reported original primary data and were written in English.

Abstracts, unpublished data, and letters to the editor were also evaluated. Articles meeting the inclusion criteria after a title and abstract screen underwent full article review. The reference lists of all selected papers were further reviewed to identify additional potentially relevant articles.

The detailed search strategies are provided in supplementary **Table S1**. Results are reported according to the Preferred Reporting Items for Systematic Reviews and Meta-analyses (PRISMA) guidelines.^13^

### Data extraction

A full-text review was performed on the included articles and the following data were collected: total number of patients, number of patients affected with HS, the total number of female patients (when available), total number of females with HS, total number of male patients (when available), total number of males with HS, diagnosis method (self-reported or clinician diagnosis). In addition to these data points, we also collected the study author, date, study period, and country.

### Risk of Bias and Quality Assessment

Studies were assessed for quality with the Newcastle-Ottawa Scale (NOS).^14^ An adapted version of the NOS was used to assess cross-sectional studies. The tool includes three domains: selection, comparability, and outcome/exposure. The threshold for good quality was a minimum score of 7 across all three domains. Studies with an NOS score ≥ 7 were included in the study.

### Obesity prevalence rates

The average adult Body Mass Index (BMI) scores and obesity prevalence rates in 2016 for each country were collected from the Our World in Data database published by the University of Oxford.^15^. BMI and obesity prevalence rates are reported separately by sex. A national obesity prevalence and BMI were calculated by averaging the reported female and male obesity prevalence and BMI rates, respectively.

### Smoking prevalence rates

The average adult tobacco use prevalence rates in 2016 for each country were collected from the Our World in Data database published by the University of Oxford.^15^. A national smoking prevalence was calculated by averaging the reported female and male smoking prevalence rates.

### Statistical analysis

Statistical analyses were performed using *R* version 3.6.1 (*R* Foundation for Statistical Computing). Meta-analysis of prevalence using only inverse variance methods places undue weight on studies with prevalence close to 0 or 1. Transformation of proportions helps to minimize these effects through stabilization of variances.^16^ Prevalence estimates underwent transformation using the double-arcsine method followed by an inverse-variance weighted random-effects metaanalysis using the DerSimonian and Laird method.^17^ The double-arcsine method was chosen over the logit method as it is better able to stabilize variances for proportions close to 0 or 1.^16^ We obtained a pooled HS prevalence with a 95% confidence interval (CI).

Heterogeneity was assessed using I^2^, which describes the total variance between studies due to heterogeneity of included studies.^18^

## Results

### Study selection

A literature search returned 932 potentially relevant studies of which 12 studies^19–30^ were included in the meta-analysis (**Fig. 1, Table 1**): three cohort and nine cross-sectional studies. These 12 studies comprised a combined study population of 53,794,009 across 9 different countries. Eleven studies^19–28,30^ reported the prevalence and three studies reported the incidence.^23,24,29^ Most studies examined patients of all ages. Seven of the eleven studies relied on a clinician diagnosis of HS while the remaining four studies included self-reported HS. All studies passed the NOS quality assessment threshold of ≥7 points (**Table S2**).

**Table 1.**
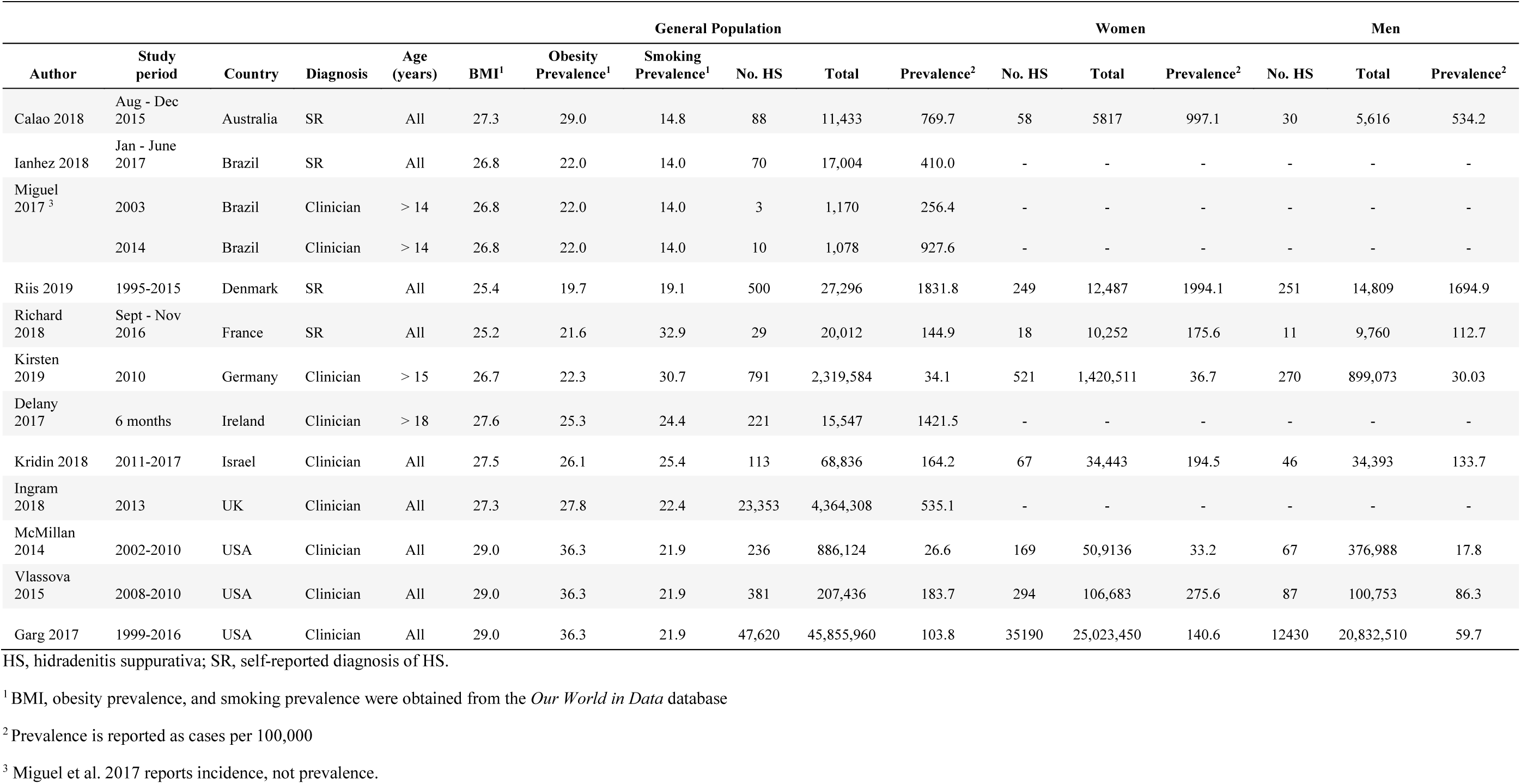
Summary of the included studies

**Figure 1.**
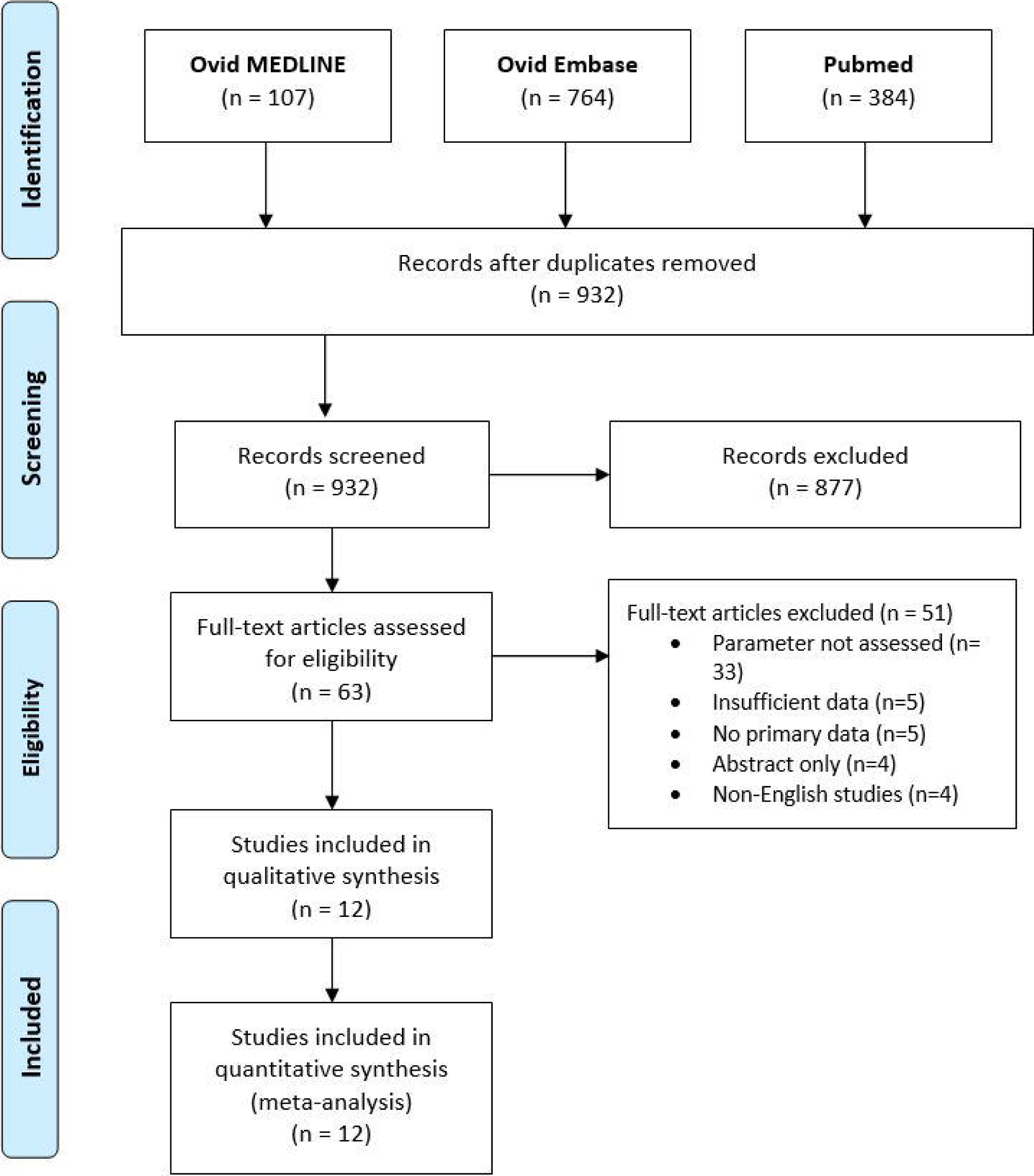
Flowchart of data selection process.

### Incidence of HS

Three out of eleven studies reported incidence of HS.^23,24,29^ Among the three studies, there was an incidence rate for each year from 1988 to 2015. Ingram et al.^23^ reported a mean annual incidence rate for physician-diagnosed cases from 1996 to 2013 of 28.3 per 100 000 person-years in the United kingdom. Miguel et al.^29^ reports an increase in incidence of HS in Brazil from 0.3% in 2003 to 0.9% in 2014. Kirsten et al.^24^ reports the lowest incidence rate of HS in Germany of 0.03% in 2015. Incidence rates are plotted in supplementary **Fig. S1**. A pooled average annual incidence rate for 1988-2015 was found to be 28.5 cases/100,000 (95% 26.8 – 30.1) (**Table 2**).

**Table 2.** Meta-analysis of proportion results

| Group | Pooled proportion | 95% CI – Lower limit | 95% CI – Upper limit | I<sup>2</sup> | P-value |
| --- | --- | --- | --- | --- | --- |
| **Prevalence** |  |  |  |  |  |
| **All** | 0.36% | 0.21% | 0.56% | 99.97% | 0.000001 |
| **Females** | 0.30% | 0.20% | 0.43% | 99.78% | 0.000001 |
| **Males** | 0.18% | 0.12% | 0.25% | 99.37% | 0.000001 |
| **Normal BMI** | 0.06% | 0.01% | 0.15% | 99.94% | 0.000001 |
| **Overweight BMI (>25)** | 0.47% | 0.20% | 0.85% | 99.93% | 0.000001 |
| **BMI <27** | 0.11% | 0.05% | 0.18% | 99.83% | 0.000001 |
| **BMI >27** | 0.58% | 0.22% | 1.11% | 99.94% | 0.000001 |
| **Obesity <27%** | 0.21% | 0.14% | 0.29% | 99.75% | 0.000001 |
| **Obesity >27%** | 0.59% | 0.18% | 1.24% | 99.96% | 0.000001 |
| **Smoking <22%** | 0.40% | 0.14% | 0.81% | 99.97% | 0.000001 |
| **Smoking >22%** | 0.31% | 0.10% | 0.62% | 98.61% | 0.000001 |
| **Age ≤ 40** | 0.16% | 0.11% | 0.22% | 99.50% | 0.000001 |
| **Age > 40** | 0.09% | 0.05% | 0.15% | 99.90% | 0.000001 |
| **Incidence** |  |  |  |  |  |
| **Average annual incidence** | 0.03% | 0.03% | 0.03% | 93.64% | 0.000001 |

CI, confidence interval; BMI, body mass index.

### Prevalence of HS

A prevalence estimate of HS was found for each study and presented with its 95% CI (**Fig. 2**). The general population estimates ranged between 0.03% and 1.80% and a pooled proportion was found to be 0.36% (95% CI 0.21 – 0.55). When the studies were divided by diagnostic method, prevalence of self-reported HS was found to be 0.66% (95% CI 0.33 – 1.09) while physician-diagnosed HS was found to be 0.24% (95% CI 0.10 – 0.44). Eight studies reported prevalence including patients’ sex (27,122,779 females and 22,273,902 males).^19,21,24–28,30^ The pooled proportions were 0.30% (95% CI 0.21 – 0.41) and 0.19% (95% CI 0.12 – 0.27) for women and men, respectively (**Fig. 3**).

**Figure 2.**
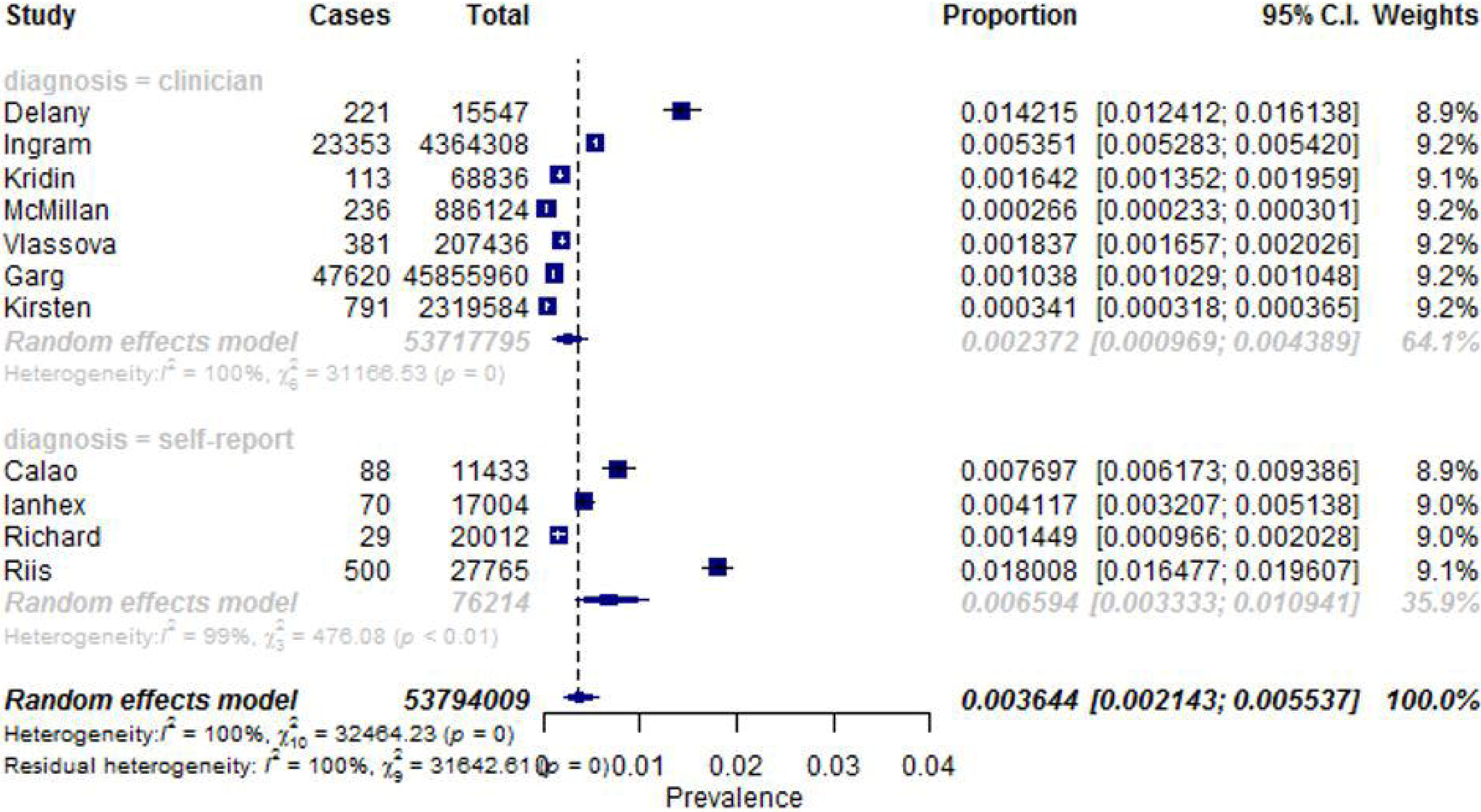
Forest plots of the prevalence of HS stratified by the method of diagnosis (self-reported versus as diagnosed by a physician).

**Figure 3.**
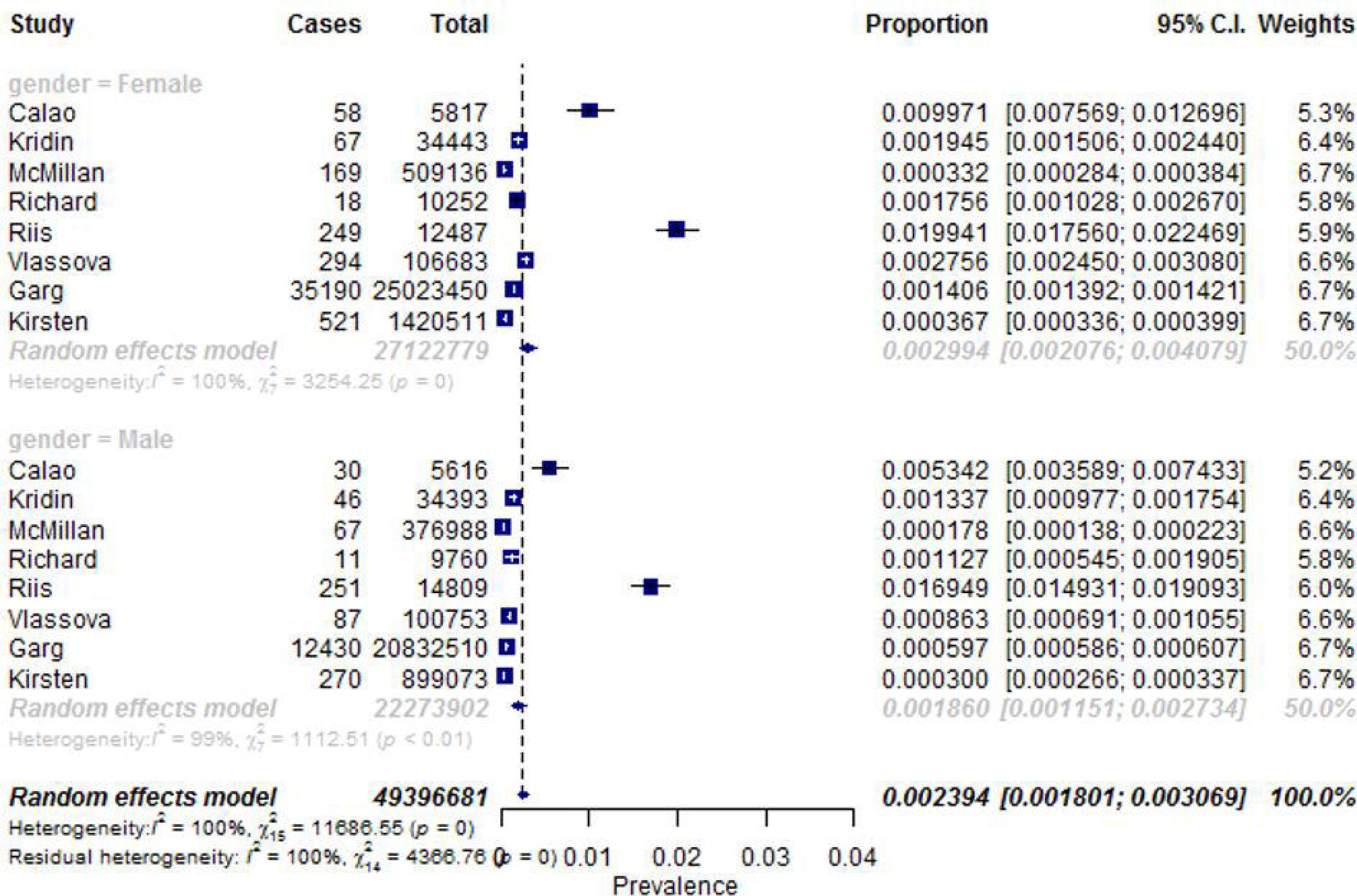
Forest plots of the prevalence of HS stratified by gender.

### Impact of patient’s age and risk factors (obesity, smoking) on HS prevalence

Five of the eleven studies reported prevalence by age group.^19,21,24,26,28^ Overall, HS prevalence was evenly distributed across age groups apart from a few outliers (**Fig. S2)**. HS prevalence was found to be 0.16% (95% CI 0.11 - 0.22) in populations aged ≤ 40 years. In populations over 40, HS prevalence was wound to be 0.09% (95% CI 0.05 - 0.15).

Two countries, Denmark and France, had a healthy national BMI of ≤ 25. The pooled prevalence of HS between these two countries was 0.06% (95% CI 0.01 - 0.15) (**Table 2**). The remaining studies had national BMI rates over 25, putting them in the overweight range. The pooled prevalence of HS of these remaining studies was 0.47% (95% CI 0.20 - 0.85). The average national body mass index (BMI) was 27 among the countries analysed. The pooled HS prevalence was found to be 0.11% (95% CI 0.05 – 0.18) and 0.58% (95% CI 0.22 - 1.11) in populations with an average BMI below and above 27, respectively (**Table 2**). When the national BMI of each study and estimated prevalence of HS were plotted into a curve (**Fig. S3**), no significant association was observed between higher prevalence of HS and BMI.

The average national obesity prevalence among the countries analysed was 27%. The pooled HS prevalence was found to be 0.21% (95% CI 0.14 – 0.29) and 0.59% (95% CI 0.18 - 1.24) in populations with an average obesity prevalence below and above 27%, respectively (**Table 2**). When the national obesity prevalence rate of each study and the estimated prevalence of HS were plotted into a curve (**Fig. S4**), no significant association was observed between higher prevalence of HS and obesity prevalence.

The average national smoking prevalence among the countries analysed was 22%. HS prevalence was found to be 0.40% (0.14 - 0.81) and 0.31% (0.10 - 0.62) in populations with an average smoking prevalence below and above 22%, respectively. A plot of the national smoking prevalence rate of each country and the estimated prevalence of HS (**Fig. S5**) showed no significant association between higher prevalence of HS and tobacco use.

### Study heterogeneity

All meta-analyses, including sub-group analyses, showed high heterogeneity with I^2^ values greater than 93% and statistically significant p-values <0.001 (**Table 2**).

## Discussion

To our knowledge, this is the first meta-analysis reporting the world-wide prevalence and incidence of HS. We found that 0.36% of the general population is affected by HS and an annual average incidence rate of 28.5 cases/100,000 over the period 1988-2015.

The diagnostic method had a great impact on reported prevalence of HS. In the studies utilizing self-reporting the prevalence was the double of the prevalence reported in the clinician-diagnosed cases. As HS is a largely clinical diagnosis, the sensitivity and specificity of a diagnosis will depend on the clinician’s experience and background. Of the seven studies reporting clinician-diagnosed HS, only two were diagnosed by a dermatologist. HS may be under-recognized in primary care practices, contributing to the lower prevalence in the clinician-diagnosed group. However, recent studies have reported that general practitioners’ recognition of HS is comparable to dermatologists.^31^ Furthermore, HS prevalence depends on patients reporting their symptoms. HS patients may be less likely to present for medical care due to the embarrassment and social stigma associated with the diagnosis.^32^

Previous studies reported a female predominance of HS ranging between 2:1 to 5:1.^33,34^ HS seems to be influenced by sex hormones because it often shows premenstrual flares, improve during pregnancy and may benefits from anti-androgenic therapy.^35^ Our study confirmed that HS affects women more than men, with a ratio of 3:2.

HS is reported to be associated with obesity and thought to be due to larger intertriginous folds and increased mechanical friction causing injury to follicles progressing to follicular occlusion and skin irritation.^36^ Though our study did find a slightly higher prevalence of HS in populations with a higher BMI and rate of obesity, those correlations were not statistically significant. The included populations had similar average BMI rates which could be the reason for lack of significant association. Further studies investigating the association of weight and HS prevalence, controlling for gender, are needed.

Although previous studies have reported a positive association between HS and smoking^37^ our study did not find a higher prevalence of HS in populations with a greater percentage of smokers.

This study is subject to limitations. All meta-analyses, including sub-group analyses, showed a high statistical heterogeneity (I^2^ >98%) suggesting that the differences in prevalence estimates between studies are not due to random chance but may be attributed to the already mentioned differences in diagnostic methods, awareness of HS, and publication bias. Furthermore, the associations between HS and risk factors such as obesity and smoking prevalence were analysed using national averages of obesity, BMI, and percentage of smokers. Those values do not necessarily reflect the prevalence of obesity and smoking in the HS population. Finally, the majority of the countries included in this meta-analysis have predominantly Caucasian populations. As evidence suggests a higher prevalence in patients with skin of color^38,39^ this study could be underestimating prevalence in other parts of the world.

## Data Availability

Included in Table 1

